# The ETKA study - Impact of early therapy termination on treated phonemes in conversational speech tasks in children aged 5 to 6 years with phonologically delayed development: protocol for a multicenter randomized controlled trial

**DOI:** 10.1101/2022.04.14.22273887

**Authors:** Doris Detter-Biesl, Bernhard Schwartz

## Abstract

**Background:** Children with speech and language disorders frequently display speech sound disorders, the treatment of which improves children’s ability to communicate. However, there is a significant gap between those therapy capacities needed and those provided in Austria. In addition, as the effect of early therapy termination has not yet been systematically evaluated, the aim of this project is to investigate the influence of an early therapy termination on phonological processes in children aged 5 to 6 years with phonologically delayed development.

**Methods/Design:** Forty 5- to 6-year-old children, currently participating in speech therapies in the Upper Austrian region will participate in this multicenter randomized controlled trial. The participants will be randomly allocated to either the intervention group (early therapy termination) or the control group (uninterrupted ongoing therapy) by envelop randomization. Phoneme accuracy in spontaneous speech determined during 15 minutes of conversational sampling and phoneme accuracy for single-word production determined with the PLAKKS assessment will be used to assess therapy efficacy. The results from both groups will be compared regarding the correct use of the target phoneme/consonant cluster in conversational speech situation.

**Discussion:** This study will explore the effect of early therapy termination on speech sound disorders in 5- to 6-yearold children. If an early termination exhibits non-inferiority to the current ongoing therapy, therapy resources could be used more effectively. Subsequently, earlier therapy starts for more children could be facilitated and negative consequences of delayed therapies could be counteracted.

**Ethics and dissemination:** The study was approved by an institutional review board (ID: A-2019-035). Dissemination will be achieved through peer-review conferences and journals.

**Trial registration:** The study is registered at ClinicalTrials.gov (Identifier: NCT03972891, Registered 4 June 2019, https://www.clinicaltrials.gov/ct2/show/NCT03972891).

## Introduction

Many children with speech and language disorders have speech sound disorders. The worldwide prevalence ranges between 2 and 25 %[1], with 16 % in German-speaking countries in 3- to 8-year-old children[2]. Speech sound disorders (SSD) are defined by a delayed acquisition of developmentally appropriate speech sounds[3] and are characterized by high frequency errors relative to the child’s age peers[4]. This reduces the intelligibility of speech and can have a negative impact on the ability of communication.

The terms articulation disorders, phonologically delayed disorders, phonological disorders, expressive phonological disorders or deficient output phonology are used synonymously in literature even though they essentially describe different problems and therefore cannot be applied to describe homogeneous groups[3]. For instance, several groups can be distinguished, such as articulation disorders, delayed phonological development, consequent phonological disorders and inconsequent phonological disorders[5]. In particular, speech sound disorders can have an impact on the development of literacy and place the child at risk of developing dyslexia when SSD persists beyond the age of school entry[3, 4, 6–9]. 4- to 5-year-old children with communication impairment have significant poorer reading and writing skills as well as report more bulling or poor peer relationships compared to 7- to 9-year-old peers[10].

Early detection of speech and language disorders and a timely delivery of appropriate measures/intervention in the event of disorders can prevent the secondary consequences of speech sound disorders, with the critical period for rapid service being the final two years before school entry[4]. Therefore, the goal of speech therapy is to improve speech intelligibility and to maximize the child’s communication skills for social and educational participation[11].

Existing therapy concepts differ both in theoretical background and in resulting procedures. This approach is articulatorily/motorically oriented and/or phonologically/linguistically oriented[12]. Phonological intervention is an improvement of the speech sound system, aiming at the generalization of taught and untaught phonemes and phonological structures[13, 14]. In phonological treatment children should learn the relevant characteristics of the phonology of ambient language. The selection of the phoneme targets depends on how the targets are expected to affect the child’s larger sound system. Usually, in phonological treatment more than one phoneme and meaningful words are used in therapy. In contrast to the phonologically-based intervention, the articulatory-oriented treatment focusses on the correction of individual phonemes directed at correctly isolated speech sounds[13, 15]. The choice of phoneme targets in the articulatory-oriented therapy depends on the development order of acquisition, stimulability of the isolated sounds and the effect on intelligibility. Usually, at the beginning of the articulatory therapy, there is an ear training and consequently the clinical interventions are considered to improve the children’s skills to build the target speech sounds. The length and complexity of the utterances are gradually increased until the child uses the speech sounds in conversational speech[13].

A key factor for the effectiveness of the different treatment approaches is the outcome to be achieved[16]. Particularly, due to the heterogeneity of children with speech sound disorders, different interventions are effective for each child and a variety of approaches are effective for similar problems[17, 18]. Therapeutic measures should be adapted to the needs of each child and oriented towards the international classification of functionality (ICF). Furthermore, the clinical decisions regarding assessments and intervention should be based on the evidence-based practice (EBP)[19]. According to the latest findings, there is no tailor-made intervention approach that can be used exclusively.

The quality and quantity of the intervention (therapy intensity) in a certain period of time are widely discussed [19–21]. The optimal therapy intensity to achieve a function gain on speech intelligibility cannot be determined and there is an ongoing discussion on the optimal duration of the intervention and the dose frequency[19–22].

While in phonological-oriented therapy concepts short periods of intervention with a higher intensity (more than one time per week) are reported, children in articulation therapy are guided clinically to produce the respective target phoneme in isolation aiming at continuous speech in a longer therapy phase (usually one time per week)[13]. There are indications that shorter therapy periods with increased therapy intensity (three times a week) may be more effective than longer therapy phases with fewer therapy units (e.g. one time per week)[4, 19, 21]. Interval therapy (i.e. therapy phases followed by therapy breaks) was found to be mostly effective when considering a connection with higher treatment intensity[20, 22].

Therapy intensity relies on three factors: a) number of properly implemented teaching episodes per session (dose); b) task or activity that are delivered within the teaching episodes (dose form) and c) the number of sessions per therapy phase (dose frequency) [19, 20, 23]. Nevertheless, the estimation of therapy dose measurement poses difficulties (i.e. each therapy unit would have to be recorded and analyzed in order to determine the exact dose). Measuring the dose within a therapy unit is difficult and depends on qualitative factors such as the therapist’s professional experience. The form of the dose depends, among other factors, on the therapist’s knowledge and the child’s abilities. The dose frequency as well the total duration of the intervention – the period between the first contact and the end of the treatment[18] –, can be quantitatively measured but depends on the respective prescription mode, expert opinions and on factors that can hardly be influenced (illnesses, holidays, etc.)[22]. In addition to the discussion on therapy intensity, the total amount of therapy hours cannot be generalized for each child. Twelve to twenty hours of intervention as well as parental inclusion have shown improvements in the functional capacity of the linguistic output[17]. It remains to be seen which treatment, which treatment intensity and thus which duration of the treatment is required to achieve functional gains in conversational speech[4, 20].

Unfortunately, due to the high demand for speech and language therapy and the shortage of therapists in the suburban and rural areas, many children are on the waiting list for speech and language therapies. In some parts of this country, recent surveys exhibited an average delayed start of speech and language therapies between from 3 to 6 months, which in some cases can exceed more than one year[24, 25].

### Study objectives

There is an ongoing discussion about the frequency and duration of speech and language therapy. Due to this, the criteria for therapy breaks, intervals and termination are based on the clinical experience rather than clinical evidence. Hence, this study design was developed to answer the following primary research question:

Do early therapy terminations in children aged 5 – 6 years with phonologically delayed development significantly impact the use of the target sound / target consonant cluster in spontaneous speech if a threshold value of 70 – 80 % correct use of the target sound is reached before early therapy termination?

### Hypothesis

We claim in our hypothesis that early therapy termination after 70 – 80 % correct use of target sound is non-inferior to a continuous therapy on conversational speech tasks.

## Material and Methods

### Aims

As persistent SSD beyond primary school entrance age can have negative long-term consequences on social, academic and professional development, waiting times for speech and language therapy should not exceed 2 months[4]. However, throughout Austria there is a shortage of 127,000 speech and language therapy places for children and young adults[24], and it is necessary to find appropriate solutions for this problem. One approach could be the early termination of therapies after a predefined threshold. Hence, the aim of this randomized controlled trial was to investigate the efficiency of an early-terminated (i.e. after 70 – 80 % correct use of the target sound) speech therapy.

### Study design

ETKA is a multicenter randomized controlled parallel two arm trial comparing early therapy termination (ETT – intervention) with an ongoing therapy (OT – control condition). Children already undergoing speech therapy will be recruited by speech and language therapists of two independent institutions located in all districts in Upper Austria. Primary assessments will be performed at baseline (t_0_) and after 12 weeks (t_1_). In addition, follow-up assessment will be performed after school entry, prior to the introduction of the grapheme of the target sound / sounds (t_2_) and 1 – 2 weeks after the introduction of the grapheme of the target sound / sounds (t_3_). The SPIRIT study schedule is illustrated in Fig 1.

**Fig 1.**
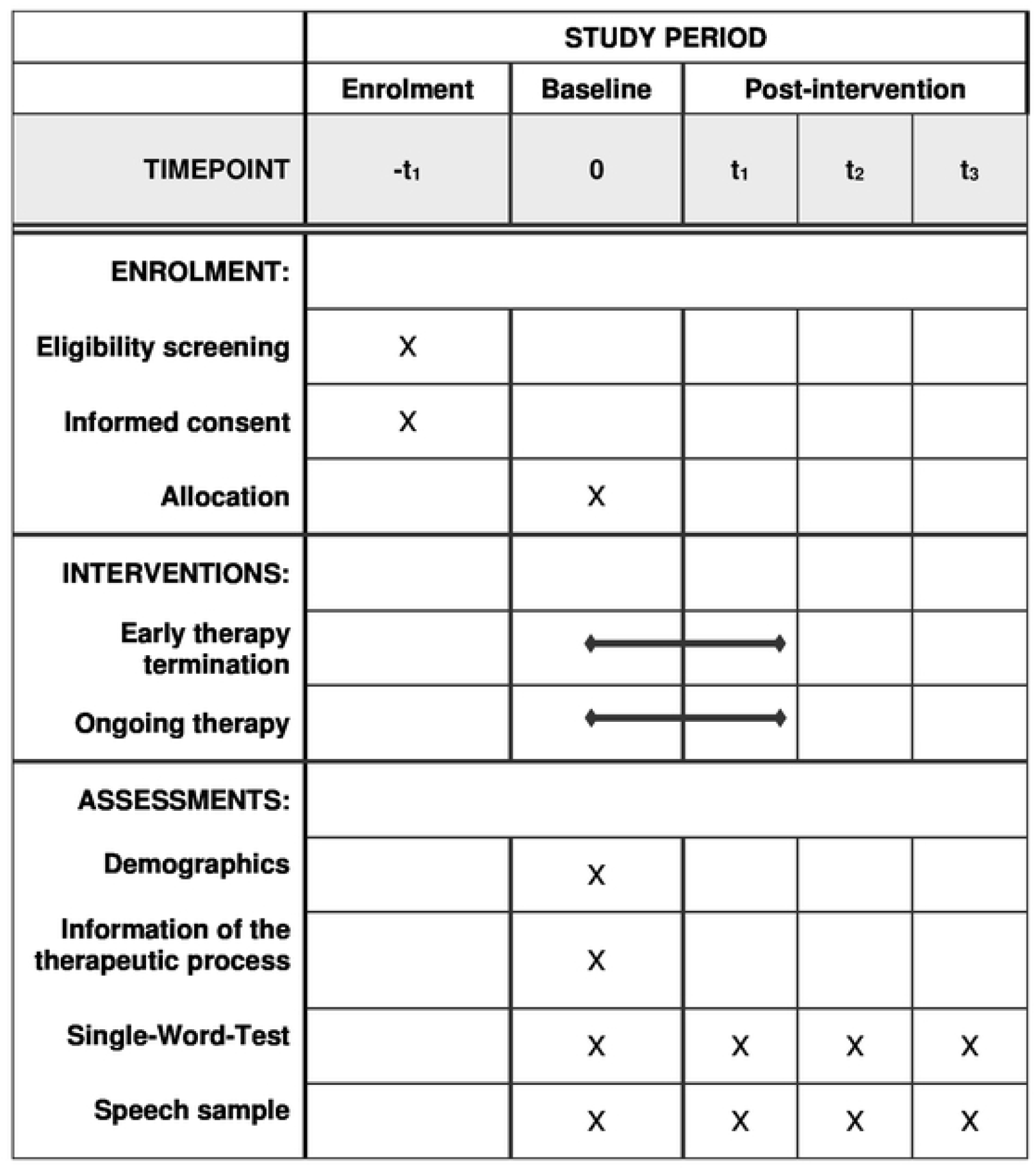
SPIRIT study schedule of enrolment, interventions, and assessments.

### Participants selection

The study population consists exclusively of children who are already undergoing speech therapy. Children between the ages of 5 and 6 of all sexes and ethnicities will be represented in the collective.

### Recruitment centers

The participants will be recruited by speech and language therapists (SLP) of two independent institutions, *Caritas für Kinder und Jugendliche* and *Volkshilfe GSD GmbH*. The therapy institutions are located in all districts in Upper Austria and children are already undergoing speech therapy treatment. Eligible children - not receiving financial benefits for study participation - will be enrolled in the first half of 2022.

The completion of the study enrolment is expected by May 2023.

### Subject inclusion criteria

#### Children are included under the following criteria

a. phonological delay of one or two of these phonological processes: palatal fronting /sch/ to /s/ or /ch/ to /s/, velar fronting /k g/ to /t d/ or contact assimilation /dr tr/ to /gr kr/,
b. correct use of the treated phoneme / consonant cluster in conversational speech situations during therapy from 70 – 80 %,
c. the phonological process has not yet been treated by another speech therapist,
d. informed consent (children and legal guardians)
e. therapy frequency on a regular weekly basis, but at least once every two weeks,
f. physiologically developed prescriptive skills,
g. good knowledge of German.

### Subject exclusion criteria

#### Children are excluded under the following criteria

a. children younger than 5 and older than 6 years,
b. correct use of the treated phoneme / consonant cluster in conversational speech situations during therapy to less than 70 %,
c. other speech sound disorders: consistent or inconsistent phonological disorders or isolated articulation disorders or childhood apraxia of speech
d. myofunctional disorders
e. speech and language disorders in the context of comorbidities,
f. deficits in prescriptive skills.

### Randomization and blinding

After the baseline assessment is completed, the children are randomly allocated to either the intervention group (ETT) or the control group (OT) by envelopes. Each envelope (i.e. 20 blue and 20 white envelopes) was previously filled with the group assessment (“early therapy termination” or “continuous therapy”) and signed by an independent biometrician. White envelopes will only be used when all blue envelopes are opened to ensure that 20 group-balanced children can be recruited for interims analysis. The non-transparent envelope is randomly drawn and opened by the therapist. The randomized result, the name of the child and the date of the assignment will be documented in the case report form (CRF).

For children enrolled in the ETT Group, therapy will be terminated early and children randomized in the OT Group will receive ongoing therapy until more than 90 % of the target phoneme / target consonant cluster can be used correctly in spontaneous speech situations (for max. 12 weeks). Due to the intervention type (i.e. early termination of on ongoing therapy) this study is planned as an open-label study. Hence, study participants (children and their caregivers), the speech and language pathologists, who administered the clinical testing and treatment, and the study leader are not blinded to the intervention.

### Sample size

The sample calculation was carried out using the program G* Power 3.1.9.2. The sample calculation is based on the correctness of the target phoneme in spontaneous speech situations in the control group from 96 ± 3 %, a limit of 93.12 ± 3 % for the equivalent consideration. It is also based on a conservative sample calculation (not normally distributed, bilaterally) considering the alpha-value of 5 %, the beta-value of 20 %, a balanced group ratio und a drop-out rate of about 10 %. Based on these calculation assumptions, 40 children must be included in the study in order to achieve a sufficient statistical power above 0.80.

### Study groups

The group allocation starts as soon as 70 – 80 % of the target sound / target consonant cluster can be used correctly in conversational speech during therapy tasks.

#### Intervention group (early termination of therapy)

Within the intervention group, the speech and language therapy will be stopped. This also includes a stop of providing language-specific exercises by the caregivers. Nevertheless, language-specific strategies in everyday life can be applied (e.g. modelling techniques).

#### Control group (ongoing therapy)

Within the control group, children will continue to receive ongoing speech and language therapy until more than 90 % of the target sound / target consonant cluster can be used correctly in conversational speech situation (for max. 12 weeks).

### Outcome and study assessments

As illustrated in Fig 1 the outcome measures will be assessed at baseline(t_0_): as soon as 70 – 80 % of the target sound / target consonant cluster can be used correctly in spontaneous speech tasks, and 12 weeks after baseline (t_1_). The follow-up measurements will be performed after school entry prior (t_2_) and after (t_3_) the introduction of the grapheme of interest.

### Outcome measures

The measurements are performed by speech and language therapists who have at least two years of experience in the field of children’s speech and language therapy.

As data collection influences articulation accuracy and articulation errors[26], two different measuring methods (i.e. picture-word articulation tests and continuous speech samples) will be used. The same materials and situation pictures for all test times allow for a direct comparison.

#### Single-word test

The articulation accuracy for single-word will be determined by the standardized single-word test “*Psycholinguistische Analyse kindlicher Aussprachestörungen* (PLAKKS)”[27]. This assessment is usually used to determine speech sound disorders in children between 2.6 and 5.11 years. PLAKKS[27] is commonly available for all therapists and it consists of 99 colored pictures that have to be named by the test subjects.

#### Conversational speech sample

Conversational sampling is a common method of measurement to describe the phoneme accuracy in conversational speech[28, 29]. In this study the length of the speech sample was set to 15 minutes to ensure a recording (recorded with a digital voice recorder) with a sufficient amount of words (at least 100 words) and target sound / target consonant cluster.

Picture description using funny situations pictures in a directed play will be used to motivate children to talk. For the sounds /sch/ and /ch/ the picture materials are called “*Such das!*”, and for /k g/ and the consonant cluster /tr dr/ it is called “*Plappersackkarten*”. In order to provide equal conditions for all children, the same pictures are selected for all children. To produce conversational speech the therapist uses a set of cues described in Rvachew & Brosseua-Laprè[20]. Each cue consists of two possible answers (yes/no answers should be avoided) and by giving suggestions and assumptions, spontaneous statements by the test subjects can be evoked.

### Data collection, management, and analysis

For analyzing the single-word-test, the speech and language therapists have to write the utterance in phonetic transcription in a protocol sheet.

Analysis of the recording of the speech samples will be carried out by the study management. The transcription of the speech samples will be done orthographically and in a broad phonetic transcription by using the International Phonetic Alphabet (IPA). The analysis will be based on the rule of the application Percentage of Consonants Correct (PCC)[30]. The percentile rank will be determined by counting the total of all target sounds produced by the child and determining the proportion that will be correctly formed. For evaluating the correct consonants the rules of the PCC[30] apply.

Several previous studies on the effectiveness of phonological therapy defined the therapy intensity, and the therapeutic method was mentioned to potentially influence the generalization of the target sounds into conversational speech[11, 17, 19, 31]. Therefore, the study authors’ query follows parameters at measurement time point t_0_: number of therapy units before enrolment, involvement of parents and information on the therapeutic process before inclusion.

#### Data validation

Validation of data is made by programmed checks of range, validity, and consistency (e.g. range checks for data values) by study members. If necessary, queries are made by the study software or an authorized person. Based on the queries the investigator can check and clarify discrepancies. After the recording of all entries and clarification of all queries, the data base will be closed upon completion of the study. An external data monitoring committee (DMC) was not formed for this study due to financial issues.

#### Data protection

All the data collected in this study may only be recorded by professionally qualified and authorized persons. All the participants will be given a unique and untraceable study number (ID) listed on a separate document. This identification list is only available to a selected circle of people, will remain in the study center after the study has finished and will not be given to the sponsor. Measurement data will only be stored in a pseudonymized way and the data security process follows the principle of good clinical practice and comply with the national data security regulations and/or the EU General Data Protection Regulation (GDPR).

### Statistical analysis

Statistical analysis will be conducted using SPSS. In addition to descriptive statistics (e.g. means, standard deviations, 95 % confidence intervals) inference statistics will be applied. For normal distributed data of variance ANOVA will be used to analyze the group-by-time interaction effect. Paired and unpaired t-tests as well as their non-parametric equivalent (i.e. Wilcoxon-Signed-Rank-test & Mann-Whitney-U-test) will further be used to analyze within-group and between-group differences, respectively. Repeated measure ANOVA and Friedman-tests (for non-parametric metric parameters) will be applied for within-group differences, too (> 2 time points). To test for normality and homogeneity of variance, Kolgomorow-Smirnov or Shapiro-Wilk-Test and Levene-test will be used. In addition, Chi-squared and McNemar-test will be applied for nominal parameters. In general, two-sided tests with an alpha risk of 0.05 a beta risk of 0.2 will be accepted. For multiple testing, alpha values will be corrected with the Bonferroni procedure.

#### Population analysis

In accordance with current recommendations by national authorities (e.g. FDA guidelines like “Non-Inferiority Clinical Trials to Establish Effectiveness - Guidance for Industry), “Intention-to-treat” (ITT) and “Per-protocol” (PP) analysis will be executed.

#### Interims analysis

An interim analysis will be executed after 20 participants completed the last assessment day. If interim analysis exhibits a significant worsening in conversational speech situation for the ETT participants and for study leavers, the conventional speech situation will be continued.

#### Missing values

For single parameters with less than 5% of missing data, multiple imputation methods will be applied. For parameters with more than 5% of missing data, all data will be analyzed as available. The number and/or relative frequency of subjects prematurely leaving the study, subjects with (major) protocol deviations (leading to exclusion from the per-protocol set) and missing values will be listed and descriptively analyzed.

### Ethics approval and consent to participate

The study was provided by the compliance with the provisions of the Helsinki Declaration ethical standards for clinical research and is in line with the rules of good scientific practice for future research projects or academic work. The study protocol (v1.01 from 21.01.2019) was approved by universities’ institutional review board (“*Ethik-und Forschungskomitee (EF) der FH Gesundheitsberufe OÖ*”) in February 2019 (A-2019-035) and was published on ClinicalTrials.gov in June 2019 (Identifier: NCT03972891). The ethics board will be informed about all substantial study protocol changes via amendments. The amendments will be signed by all the signatories to the original protocol.

All participants, the legal guardians of the children and the therapists, will be informed about the study by the study members. In addition, children, and their legal guardians (i.e. mainly the parents) will receive additional information about the study procedure by the speech therapist. While the study participants (i.e. children receiving speech therapy) will be asked for a verbal informed consent, their legal guardians will be asked to sign a written informed consent for study participation. Children’s verbal informed consent will be documented in the case report form.

### Trial status

The recruitment was planned to be started in February 2019 but has been cancelled due to the COVID-19 pandemic. Recruiting will start as soon as children will receive uninterrupted speech and language therapies again, which is expected in February 2023.

## Discussion

To date, there is no consensus in the scientific community on how often, how long, and when therapy should be terminated. Due to the insufficient speech and language therapy supply in Austria, it is particularly important to generate the optimal threshold for the termination of speech and language therapy without imposing negative effects on the success of the therapy. An evidence-based threshold at which speech therapy is effective is still unclear and has mostly been derived from descriptive studies[4] or observational studies[22]. In addition, threshold-baseline for therapy termination achieving a generalization of the learned phoneme into untreated context has not been investigated to date, or at least such studies are not known to the author despite research. However, ETKA aims at filling this research gap.

The ETKA study will determine whether early therapy termination in children between the ages of 5 and 6 with phonologically delayed development of one phonological process will lead to similar effects in the transfer of the target sound / target consonant cluster in continuous speech as uninterrupted, ongoing therapy does. Although the planned study is not based on a pilot study, assumptions on an early termination threshold (i.e. 70 – 80 % correct use of the treated phoneme in a conversational speech situation) are based on an existing interval therapy concept, called Psycholinguistic Phonologically-oriented Therapy (P.O.P.T)[32]. Within the course of this concept, improvements up to 90 % correctness of the target objective occurred after 12-week-therapy interruptions or early terminated therapy after 60 – 80 % correct implementation of the respective objective. In addition, some literature exposed that the performance level which children should reach in order to apply the correct sound to untrained words ranges between 40 and 75 % in correctness[20].

The ETKA study exhibits some limitations. Although the biometrist responsible for the statistical analysis will be blinded within this study, blinding based on the intervention type cannot be realized for the therapists and the participants. In addition, although some basic approaches for spontaneous speech assessments exist, the elicitation of spontaneous speech is not internationally standardized. Nevertheless, to reduce this performance bias, identical tasks (i.e. situation pictures and instructions) will be used in all implemented continuous speech measurements. Unfortunately, within this study statements about the cumulative intervention intensity cannot be drawn, because the number of implemented teaching episodes per session (dose) before early interruption starts cannot be estimated appropriately enough. Lastly, as each SLP can individually choose the speech sound therapy, study participants may have different therapy experience prior to the study inclusion.

Contrary to these limitations, ETKA study has several strengths, including the randomized controlled trial design, an *a priori* sample size calculation, strong inclusion criteria, identical environmental measurement conditions, and objective assessment of the primary outcome. By this stringent design, a maximum reduction of bias and sufficient statistical power, ETKA study – contrary to previous pilot studies – will be able to create enough evidence to determine if early therapy terminations can lead to similar therapy success as common therapies. If the ETKA study can show that an early therapy termination after reaching the 70 – 80 % in the generalization of the treated sounds into conversational speech does not have a negative impact on language acquisition, speech and language therapy resources could be used more effectively. Speech and language therapists could justify early therapy termination in parental counselling based on evidence and SLP would be able to treat more children with an identically number of working hours. Hence, more children on the waiting list could then obtain the service of speech and language therapy and long-term consequences of long waiting times could be reduced.

## Data Availability

No datasets were generated or analysed during the current study. All relevant data from this study will be made available on ClinicalTrials.gov upon study completion. In accordance with the General Data Protection Regulation of the European Union, personal data will be adequately protected and will not be publicly disclosed.

## Authors’ contributions

Both authors were equally responsible for writing the manuscript and contributing to the research design. While first author DDB was mainly responsible for the speech therapy content, BS led the development of this manuscript and was mainly responsible for the statistical planning.

The authors declare that they have no competing interests.

## Acknowledgement

The authors thank Barbara Kraxberger and Martina Bernegger from the non-profit-organisations „*Caritas für Kinder und Jugendliche”* and “*Volkshilfe GSD Gmbh*”, for agreeing to support the recruiting process and study assessments. They further thank Ulrike Haslinger for proofreading the article.

## Supporting information

**S2 Fig. ETKA SPIRIT study schedule of enrolment, intervention, and assessments**

**S1 File. ETKA SPIRIT checklist**

**S2 File. ETKA study protocol**

**S3 File. ETKA ethical approval**

**S4 File. ETKA project orders**

**S5 File. ETKA consent form speech and language therapists**

**S6 File. ETKA consent form legal guardian**

## Notes

Funding This study was fully supported by the University of Applied Sciences for Health Professions Upper Austria under [grant number P-2018-044, internal university fund, “ETKA”]. The funder did not play a role in neither the design of the study protocol nor in the writing of this article. Furthermore, the funder will not have any influence on data collection, management, analysis, or publication.

### Competing Interest Statement

The authors have declared no competing interest.

### Clinical Trial

NCT03972891

### Clinical Protocols

https://clinicaltrials.gov/ct2/show/NCT03972891

### Funding Statement

This study was fully supported by the University of Applied Sciences for Health Professions Upper Austria under [grant number P-2018-044, internal university fund, “ETKA”]. The funders had and will not have a role in study design, data collection and analysis, decision to publish, or preparation of the manuscript.

### Author Declarations

The study was provided by the compliance with the provisions of the Helsinki Declaration ethical standards for clinical research and is in line with the rules of good scientific practice for future research projects or academic work. The study protocol (v1.01 from 21.01.2019) was approved by universities? institutional review board ((?Ethik- und Forschungskomitee (EF) der FH Gesundheitsberufe OÖ) in February 2019 (A-2019-035) and was published on ClinicalTrials.gov in June 2019 (Identifier: NCT03972891). The ethics board will be informed about all substantial study protocol changes via amendments. The amendments will be signed by all the signatories to the original protocol. All participants, the legal guardians of the children and the therapists, will be informed about the study by the study members. In addition, children, and their legal guardians (i.e. mainly the parents) will receive additional information about the study procedure by the speech therapist. While the study participants (i.e. children receiving speech therapy) will be asked for a verbal informed consent, their legal guardians will be asked to sign a written informed consent for study participation. Children?s verbal informed consent will be documented in the case report form.

